# Spatio-temporal analysis and prediction of malaria cases using remote sensing meteorological data in Diébougou health district, Burkina Faso, 2016-2017

**DOI:** 10.1101/2021.04.02.21254768

**Authors:** Cédric S. Bationo, Jean Gaudart, Sokhna Dieng, Mady Cissoko, Paul Taconet, Boukary Ouedraogo, Anthony Somé, Issaka Zongo, Dieudonné D. Soma, Gauthier Tougri, Roch K. Dabiré, Alphonsine Koffi, Cédric Pennetier, Nicolas Moiroux

**Affiliations:** Aix Marseille Univ, INSERM, IRD, SESSTIM, UMR1252, 13005 Marseille, France; MIVEGEC, Univ. Montpellier, CNRS, IRD, Montpellier, France; Aix Marseille Univ, INSERM, IRD, SESSTIM, UMR1252, APHM, Hop Timone, BioSTIC, Biostatistic & ICT, 13005 Marseille, France; Malaria Research and Training Center—Ogobara K. Doumbo (MRTC-OKD), FMOS-FAPH, Mali-NIAID-ICER, Université des Sciences, des Techniques et des Technologies de Bamako, Bamako 1805, Mali; Direction des systèmes d’information en santé, Ministère de la santé du Burkina Faso; Institut de Recherche en Sciences de la Santé (IRSS), Bobo Dioulasso, Burkina Faso; Institut Supérieur des Sciences de la Santé, Université Nazi Boni, Bobo-Dioulasso, Burkina Faso; Programme national de lutte contre le paludisme, Ministère de la santé du Burkina Faso; Institut Pierre Richet (IPR), Institut National de Santé Publique (INSP), Bouaké, Côte d’Ivoire

**Keywords:** Geo-epidemiology, Spatial Clusters, temporal dynamics, nonlinear relationship, prediction

## Abstract

**Background:** Malaria control and prevention programs are more efficient and cost-effective when they target hotspots or select the best periods of year to implement interventions. This study aimed to identify the spatial distribution of malaria hotspots at the village level in Diébougou health district, Burkina Faso, and to model the temporal dynamics of malaria cases as a function of meteorological conditions and of the distance between villages and health centers (HCs).

**Methods:** Case data for 27 villages were collected in 13 HCs using continuous passive case detection. Meteorological data were obtained through remote sensing. Two synthetic meteorological indicators (SMIs) were created to summarize meteorological variables. Spatial hotspots were detected using the Kulldorf scanning method. A General Additive Model was used to determine the time lag between cases and SMIs and to evaluate the effect of SMIs and distance to HC on the temporal evolution of malaria cases. The multivariate model was fitted with data from the epidemic year to predict the number of cases in the following outbreak.

**Results:** Overall, the incidence rate in the area was 429.13 cases per 1,000 person-year with important spatial and temporal heterogeneities. Four spatial hotspots, involving 7 of the 27 villages, were detected, for an incidence rate of 854.02 cases per 1,000 person-year. The hotspot with the highest risk (relative risk = 4.06) consisted of a single village, with an incidence rate of 1,750.75 cases per 1,000 person-years. The multivariate analysis found greater variability in incidence between HCs than between villages linked to the same HC. The epidemic year was characterized by a major peak during the second part of the rainy season and a secondary peak during the dry-hot season. The time lag that generated the better predictions of cases was 9 weeks for SMI1 (positively correlated with precipitation variables and associated with the first peak of cases) and 16 weeks for SMI2 (positively correlated with temperature variables and associated with the secondary peak of cases). Euclidian distance to HC was not found to be a predictor of malaria cases recorded in HC. The prediction followed the overall pattern of the time series of reported cases and predicted the onset of the following outbreak with a precision of less than 3 weeks.

**Conclusions:** Our spatio-temporal analysis of malaria cases in Diébougou health district, Burkina Faso, provides a powerful prospective method for identifying and predicting high-risk areas and high-transmission periods that could be targeted in future malaria control and prevention campaigns.

## Introduction

Malaria is one of the most life-threatening diseases and poses a great socio-economic burden worldwide [1]. According to World Health Organization (WHO) estimates, the global number of malaria cases was 228 million in 2018 (95% Confidence Interval (95% CI) = 206-258 million) compared to 251 million in 2010 (95% CI = 231-278 million) and 214 million in 2015 (95% CI = 149-303 million) [1]. Although the estimated number of cases decreased by 23 million from 2010 to 2018, data for the period 2015-2018 highlight the lack of significant progress during this period. In 2018, the WHO African Region accounted for most cases (200 million or 93% of all cases), far ahead of the WHO South-East Asian region (3.4%) and the WHO Eastern Mediterranean Region (2.1%) [1]. At the time, nearly 80% of global malaria deaths were concentrated in 17 countries of the WHO African Region and in India. The WHO estimates that Burkina Faso carries about 6% of the global malaria burden [1]. Statistical data from the Ministry of Health of Burkina Faso for the year 2015 show that malaria is the main reason for consultation (45.7%), hospitalization (45.6%), and death (25.2%) in the country’s health facilities, and that pregnant women and children under 5 years are the most at risk of contracting malaria [2]. According to the Burkina Faso Malaria Indicator Survey [3], the average parasite prevalence in children under 5 years was 46% in 2014. In 2018, the number of confirmed cases reported in health facilities was 11,624,595 of which 4.14% were severe forms and 2.8% resulted in death.

The National Malaria Control Program in Burkina Faso recommends the following control strategies [4]: early case management in health facilities and at the community level, with a particular focus on children aged 3 to 59 months [5]; intermittent preventive treatment (IPT) for pregnant women; universal access to rapid diagnostic tests (RDTs) and artemisinin-based combination therapies; seasonal malaria chemoprevention (SMC) for children under 5 years; and vector control using long-lasting insecticidal nets (LLINs), indoor residual spraying (IRS), larval control, and environmental sanitation.

For strategic reasons or lack of resources, not all of these strategies are optimally implemented everywhere and all the time. Thus, in 2018, 25% of households reported not owning an LLIN (with coverage varying between 58% and 87% depending on the region) and 42% of pregnant women did not receive the recommended three doses of IPT, as reported by the Burkina Faso Malaria Indicator Survey [6].

At the same time, new tools and strategies are being developed, including administration of ivermectin, bi-impregnated nets, transmission-blocking vaccines, and conventional vaccines [7,8,9]. In Burkina Faso, the REACT project (“Insecticide resistance management in Burkina Faso and Côte d’Ivoire: A study on vector control strategies”) conducted in 2016-2018 aimed to evaluate the efficacy of strategies designed to complement LLINs, namely pirimiphos methyl-based IRS, enhanced communication, and administration of ivermectin to domestic animals.

Malaria control and prevention programs are more efficient and cost-effective when they target high-risk spatial clusters (hotspots) [10] or when they select the best times of year [11] to initiate interventions (e.g. SMC or LLIN distribution). Indeed, as numerous studies have shown, malaria incidence at the local level is heterogeneous and associated with spatio-temporal clusters [12,13,14] that are likely to maintain transmission during low-risk periods and, consequently, to increase transmission during high-risk periods [15,16,17]. Identifying these clusters can therefore help to improve the fight against malaria and to anticipate future outbreaks.

This study aimed to identify the spatial distribution of malaria hotspots at the village level in Diébougou health district, Burkina Faso, and to model the temporal dynamics of malaria cases as a function of meteorological conditions and of the Euclidean distance between villages and their corresponding health centers (HCs). Data on malaria cases were obtained through HC-based passive case detection for the 27 villages included in the REACT project.

## Materials and methods

### Study area

The study was conducted in 27 villages of Diébougou health district that were included in the REACT project. All included villages met two criteria: a population between 200 and 500 and a Euclidean distance of at least 2 km from the nearest village. A population census carried out in July 2016 by our research team found that the 27 villages were home to 7,408 inhabitants. The villages were linked to 13 HCs. Villages and HCs were geo-referenced using GPS (Figure 1).

**Figure 1:**
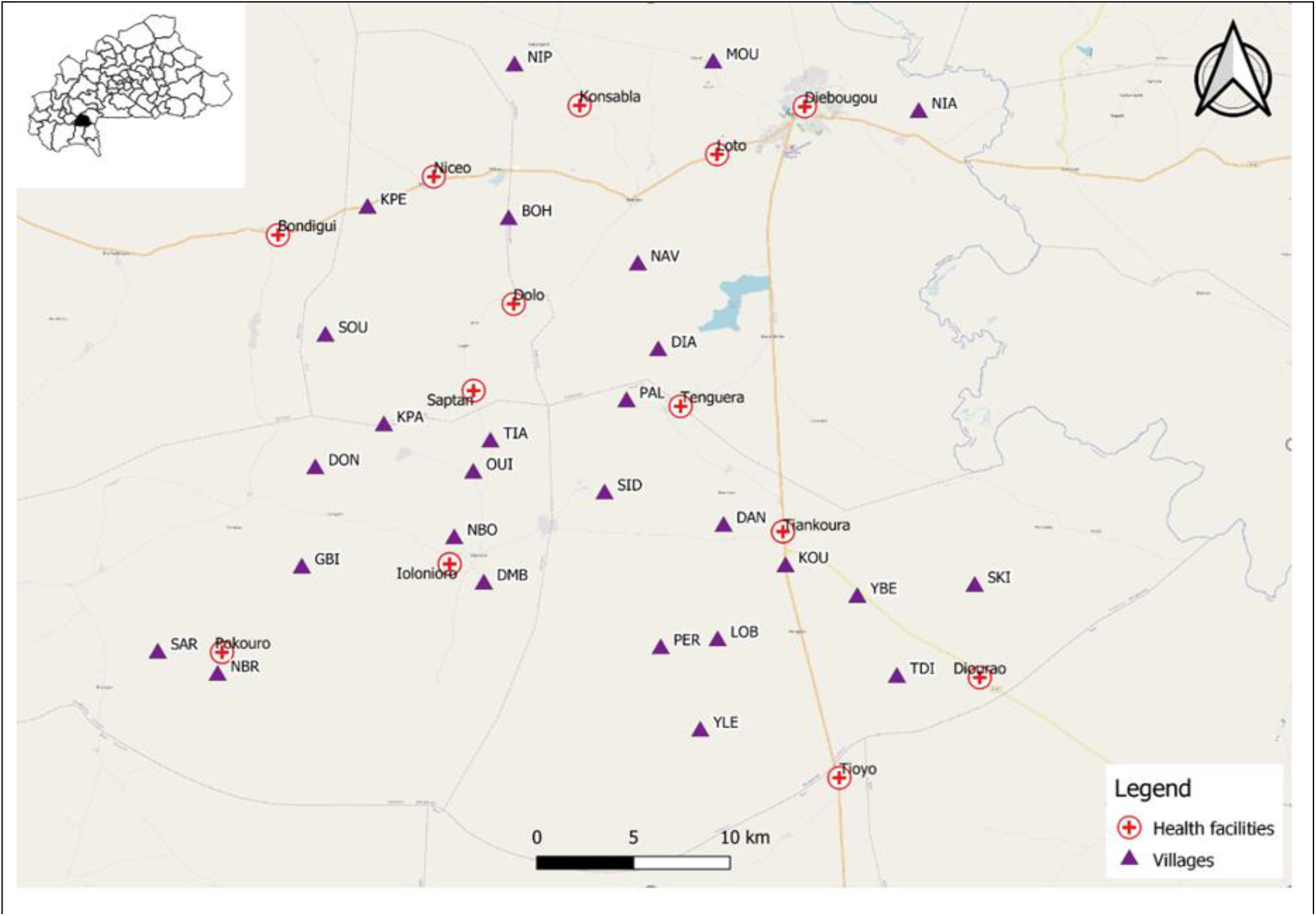
Map of the study area showing the location of villages (triangles) and health centers (red crosses). Background: OpenStreetMaps. MOU: moule; NIA: niaba; NIP: nipodja; BOH: bohero; NBR: niombripo; SAR: sarambour; NBO: niombouna; GBI: gongombiro; KPA: kpalbalo; DON: dontelo; SID: sidmoukar; DMB: dombouro; OUI: ouidiaro; TIA: tiakiro; NAV: nouvielgane; DIA: diagnon; PAL: palembera; KPE: kpedia; SOU: soussoubro; TDI: tordiero; YLE: yellela; YBE: yelbelela; SKI: sinkiro; DAN: dangbara; KOU: kouloh; LOB: lobignonao; PER: perglembiro.

Diébougou health district is located in South-Western Burkina Faso, a region characterized by a tropical climate with a dry season from October to April and a rainy season from May to September. The dry season is divided into a cold dry season lasting from December to February and a hot dry season lasting from March to April. Average daily minimum and maximum temperatures in the cold dry, hot dry, and rainy seasons are 18 and 36°C, 25 and 39°C, and 23 and 33°C, respectively. Average annual rainfall is 1,200 mm. The natural vegetation is dominated by wooded savannah dotted with clear forest gallery [18,19].

### Passive case detection

Case data for the 27 villages included in the REACT project were collected using continuous HC-based passive case detection during 2016 and the first 36 weeks of 2017, which corresponded to the period preceding the implementation of the interventions studied (i.e. pirimiphos methyl-based IRS, enhanced communication, and administration of ivermectin to domestic animals). Specifically, consultation data for village residents were retrieved from HC registries and recorded using tablets equipped with Open Data Kit collect forms. A malaria case was defined as a person who presented with fever and received a positive RDT result.

### Study period

Of the 88 weeks of data collection, 52 weeks corresponding to an epidemic year (a complete malaria epidemic) were considered for spatio-temporal analysis. The epidemic year ran from week 20 (in May) of 2016 to week 19 (in May) of 2017 (Figure 2).

**Figure 2:**
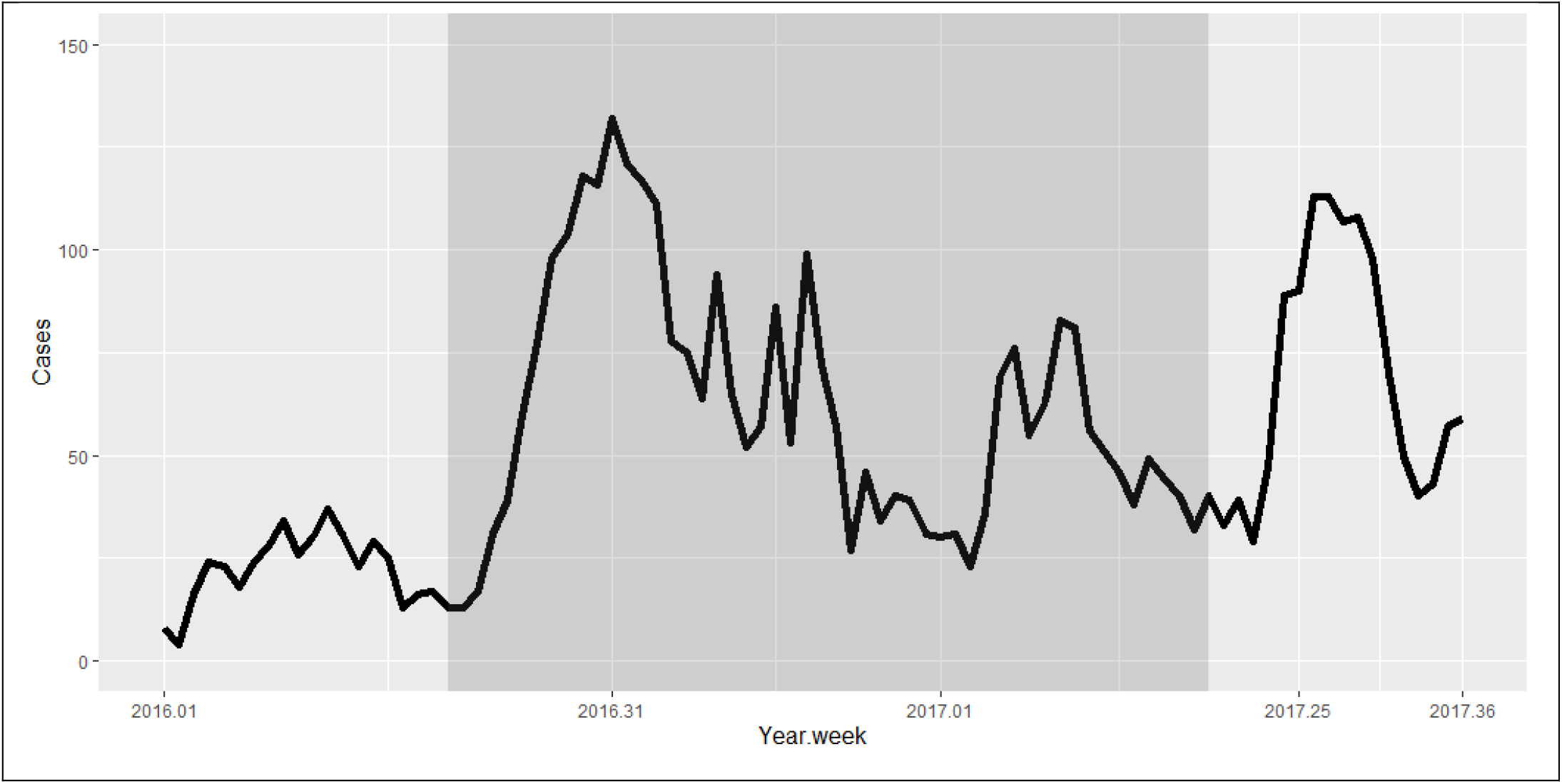
Time series of the number of malaria cases collected through passive case detection. The shaded (dark) area represents the epidemiological year considered for analysis.

### Meteorological data

The meteorological data used in this study were drawn from the Era-5 dataset [20] published by the European Centre for Medium-Range Weather Forecasts, which provides hourly estimates of several atmospheric and land parameters at a spatial resolution of 0.25° [21]. These data were aggregated into weekly counts. The meteorological variables included in the analysis were: Weekly rainfall (mm), number of rainy days per week, weekly mean of daily average temperature (°C), weekly mean of daily minimum temperature (°C), weekly mean of daily maximum temperature (°C), weekly mean of daily average wind speed (km/h), weekly mean of daily average relative humidity (%), weekly mean of daily average atmospheric pressure (hPa), weekly mean of daily average cloud cover (%), and weekly mean of daily thermal amplitude (°C) (Table 1).

**Table 1:** List of meteorological variables with their abbreviations and descriptive statistics.

| var | Mean | St. Dev. | Min | Pctl(25) | Median | Pctl(75) | Max |
| --- | --- | --- | --- | --- | --- | --- | --- |
| Tmean: weekly mean of daily average temperature (° C) | 28.7 | 2.2 | 25.7 | 26.6 | 28.8 | 30.2 | 33.9 |
| Re: number of rainy days per week | 4.8 | 2.9 | 0 | 2 | 7 | 7 | 7 |
| P: weekly mean of daily average atmospheric pressure (hPa) | 975.3 | 1.6 | 972.3 | 974.0 | 975.5 | 976.7 | 978 |
| Cl: weekly mean of daily average cloud cover (%) | 0.6 | 0.2 | 0.1 | 0.4 | 0.6 | 0.7 | 0.9 |
| W: weekly mean of daily average wind speed (km/h) | 7.6 | 3.6 | 2.8 | 4.7 | 7.2 | 9.5 | 17.4 |
| H: weekly mean of daily average relative humidity (%) | 0.5 | 0.2 | 0.1 | 0.3 | 0.6 | 0.8 | 0.8 |
| Tmax: weekly mean of daily maximum temperature (°C) | 34.4 | 3.5 | 28.6 | 31.2 | 35.4 | 36.6 | 40.7 |
| Tmin: weekly mean of daily minimum temperature (°C) | 23.3 | 1.8 | 19.1 | 22.4 | 23.0 | 24.2 | 27.0 |
| Rc: weekly rainfall (mm) | 14.4 | 18.3 | 0.0 | 0.005 | 5.0 | 24.5 | 69.8 |
| tvar: weekly mean of daily thermal amplitude (° C) | 11.1 | 3.4 | 5.9 | 7.7 | 10.6 | 14.4 | 17.1 |

*Var: Variables; St. Dev: Standard deviation; Min: Minimum; Pctl(25): First quartile; Pctl(75): Third quartile; Max: Maximum.*

To reduce the number of variables and avoid collinearity, we constructed synthetic meteorological indicators (SMIs) using a principal component analysis (PCA) of weekly meteorological variables. Principal components that met Kaiser’s criterion [22] were selected as SMIs and included in the temporal analysis.

### Spatial Analyses

Hotspots, i.e. high-risk clusters, were detected using the Kulldorf scanning method [23] with a Monte Carlo algorithm in a purely spatial analysis. The Kulldorf scanning method helps to identify spatial clusters based on geographical coordinates and to avoid the problem of multiple non-independent tests. [23]. We defined clusters as aggregates of cases with observed values higher than expected (i.e. unlikely to have been obtained by chance). The p-value (i.e. the probability, under the null hypothesis, that the expected number of cases is the same or higher than the observed number of cases) was calculated for each cluster.

Scan parameters were: elliptical window, non-overlapping clusters, maximum cluster size set at 50% of the population at risk, Monte Carlo replication number set at 999.

### Temporal analyses

#### Lagged SMI selection

Several studies have observed a lag between malaria time series and meteorological data time series [24,25,26]. In view of this, we decided to investigate the time lag (in weeks) between the time series of weekly malaria cases and the time series of SMIs. Using a generalized additive model (GAM) with a negative binomial distribution and a smoothing spline function, we modeled the time series of total malaria cases (for all villages) as a function of each SMI for time lags ranging from 1 to 30 weeks (thus generating 30 models per SMI). The GAM is an extension of the generalized linear model (GLM): while it includes random effects in the predictor like the GLM does, it can be used with nonparametric smoothing terms instead of constant parameters [27, 28, 29]. The usefulness of the GAM lies in the fact that it provides a flexible method to identify the effects of non-linear covariates in exponential family distributions and in likelihood-based methods [30,31,32]. However, instead of estimating a single parameter, the GAM provides an unspecified (non-parametric) general function that compares predicted response values to predictor values.

We compared the 30 models generated for each SMI using the unbiased risk estimator (UBRE), i.e. an unbiased estimate of the mean square error of a non-linear biased estimator. For each SMI, the time lag associated with the best model (i.e. with the lowest UBRE) was selected for the multivariate analysis.

#### Multivariate time analysis

To account for the non-linearity of the relationship between the response and predictor variables, we analyzed the time series of weekly cases reported in all villages during the epidemic year using a generalized additive mixed model (GAMM). To account for the non-independence of data from the same village or HC, we fitted this model with nested random intercepts for villages and HCs. To account for the spatial auto-correlation of the data, we used a Gaussian field with a negative exponential variogram. A first-order auto-regressive temporal auto-correlation structure was introduced to account for the temporal auto-correlation of malaria cases.

We analyzed the time series of cases using selected lagged SMIs (with a smoothing spline function) and of the Euclidean distance between villages and their corresponding HCs as predictors. For each predictor, the standardized incidence ratio (SIR) was estimated by modeling the log-transformed population as the offset.

To account for the non-linearity of the relationship between the response and predictor variables, we calculated SIRs according to the deciles of the distribution of values for each predictor. Indeed, SIRs cannot be calculated with GAMs as they are with GLMs, because when the relationship between the response and the predictor is non-linear, SIRs are not constant across the range of values of the predictor [28,32].

Lastly, we tested the multivariate model fitted with data from the epidemic year to predict the number of cases in both 2016 and 2017.

### Software and packages

Statistical analyses were performed using R software (version 3.6.1) [33]. The PCA was performed using the PCA function in the *FactoMineR* package [34]. The GAMs and the GAMM were generated using the “gam” and “gamm” functions in the *mgcv* package, respectively [30, 31, 32]. Data overdispersion was tested using the “dispersiontest” function in the *AER* package [35]. The spatial analysis was performed using SatScan™ software (version 9.6). Maps were produced using QGIS software (version 3.10) [36].

## Results

### Descriptive analysis

A total of 3,179 malaria cases were reported in HCs during the epidemic year, corresponding to an incidence of 429.13 cases per 1,000 person-years. On average, 61.13 cases per week were reported, with a peak of 132 cases in week 31 of 2016 (week 1 of August; Figure 2). The curve of cases over the epidemic year shows two peaks (Figure 2): a very pronounced peak between weeks 27 and 45 of 2016 (August to November), which accounted for 60% of cases, and a less pronounced peak between weeks 7 and 11 of 2017 (mid-February to the end of March), which accounted for 12% of cases.

### Synthetic Meteorological Indicators

The PCA conducted using Kaiser’s criterion led us to construct and retain two SMIs that explained 85.4% of the total inertia (Figure 3A).

**Figure 3:**
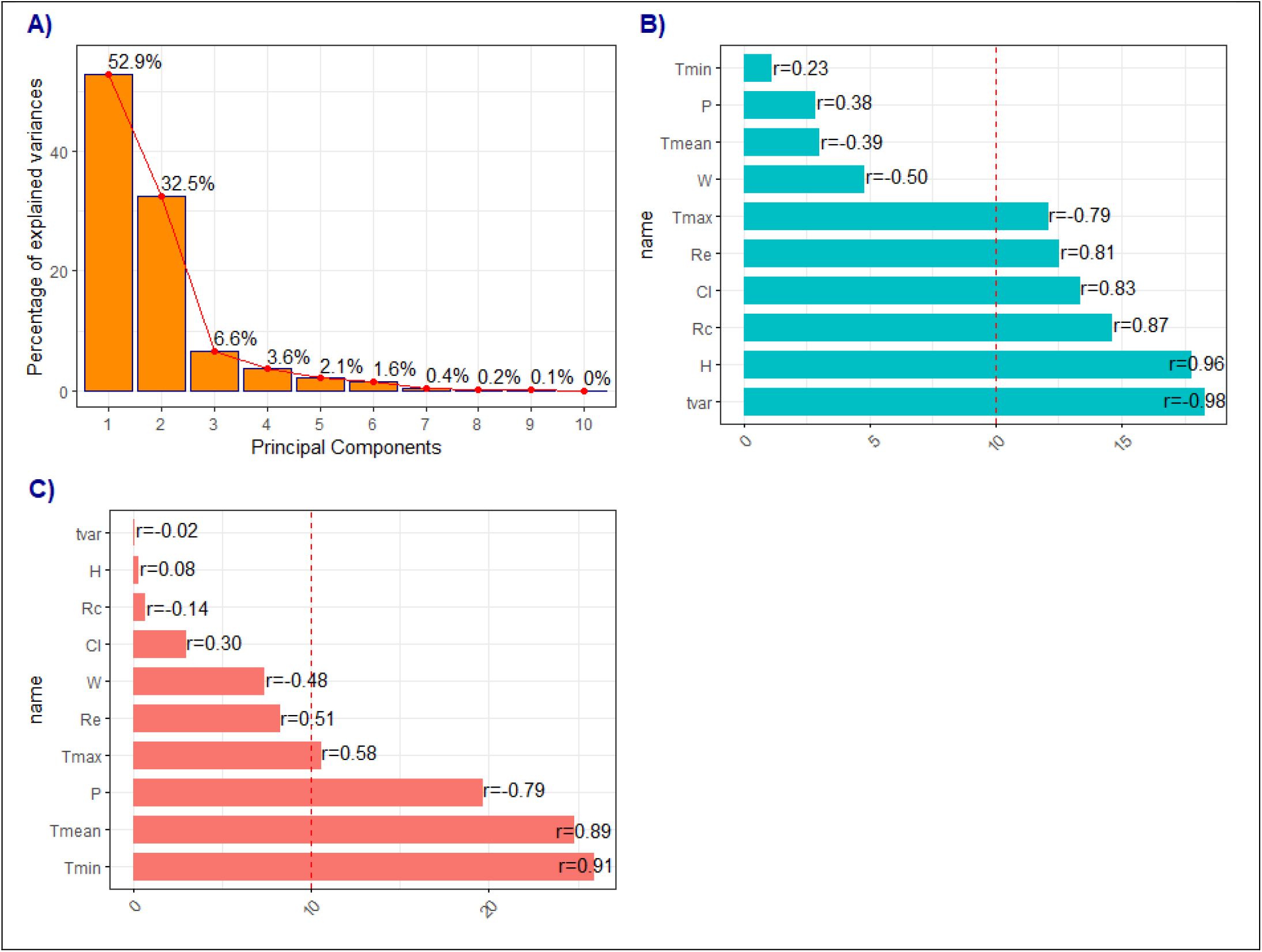
Principal Component Analysis of meteorological variables. Percentage of inertia explained by each principal component (Panel A). Contribution of meteorological variables to the first principal component (SMI1; panel B) and the second principal component (SMI2; panel C). SMI: Synthetic meteorological indicator. r: Correlation coefficient between the meteorological variable and the SMI. Abbreviations of variable names are detailed in Table 1. The dashed line represents the contribution that would have been expected if all variables had contributed equally to the SMI.

The first SMI (i.e. the first principal component) explained 52.9% of the total inertia. The variables that most contributed to this SMI, henceforth called SMI1, were mainly correlated with precipitation variables: weekly mean of daily thermal amplitude (18.24%, correlation coefficient r = −0.98), weekly mean of daily average relative humidity (17.74%, r = 0.96), weekly rainfall (14. 5%, r = 0.8), weekly mean of daily average cloud cover (13.32%, r = 0.83), number of rainy days per week (12.47%, r = 0.81), and weekly mean of daily maximum temperature (12.03%; r = −0.79) (Figure 3B). The second SMI (i.e. the second principal component) explained 32.5% of the total inertia. The variables that most contributed to this SMI, henceforth called SMI2, were mainly correlated with temperature variables: weekly mean of daily minimum temperature (25.83%; r = 0.91), weekly mean of daily average temperature (24.72%; r = 0.89), weekly mean of daily maximum temperature (10.51%; r = 0.58), and weekly mean of daily average atmospheric pressure (19.6%, r = −0.79) (Figure 3C).

The values of SMI1 were positive between late June and early October, which corresponds to the rainy season (Figure 4). The values of SMI2 were positive between mid-February and mid-June, which corresponds to the hot dry season (March - June), and between October and November, which corresponds to the transition period between the rainy season and the dry season. Both SMIs were negative throughout the cold dry season (December - mid-February) (Figure 4).

**Figure 4:**
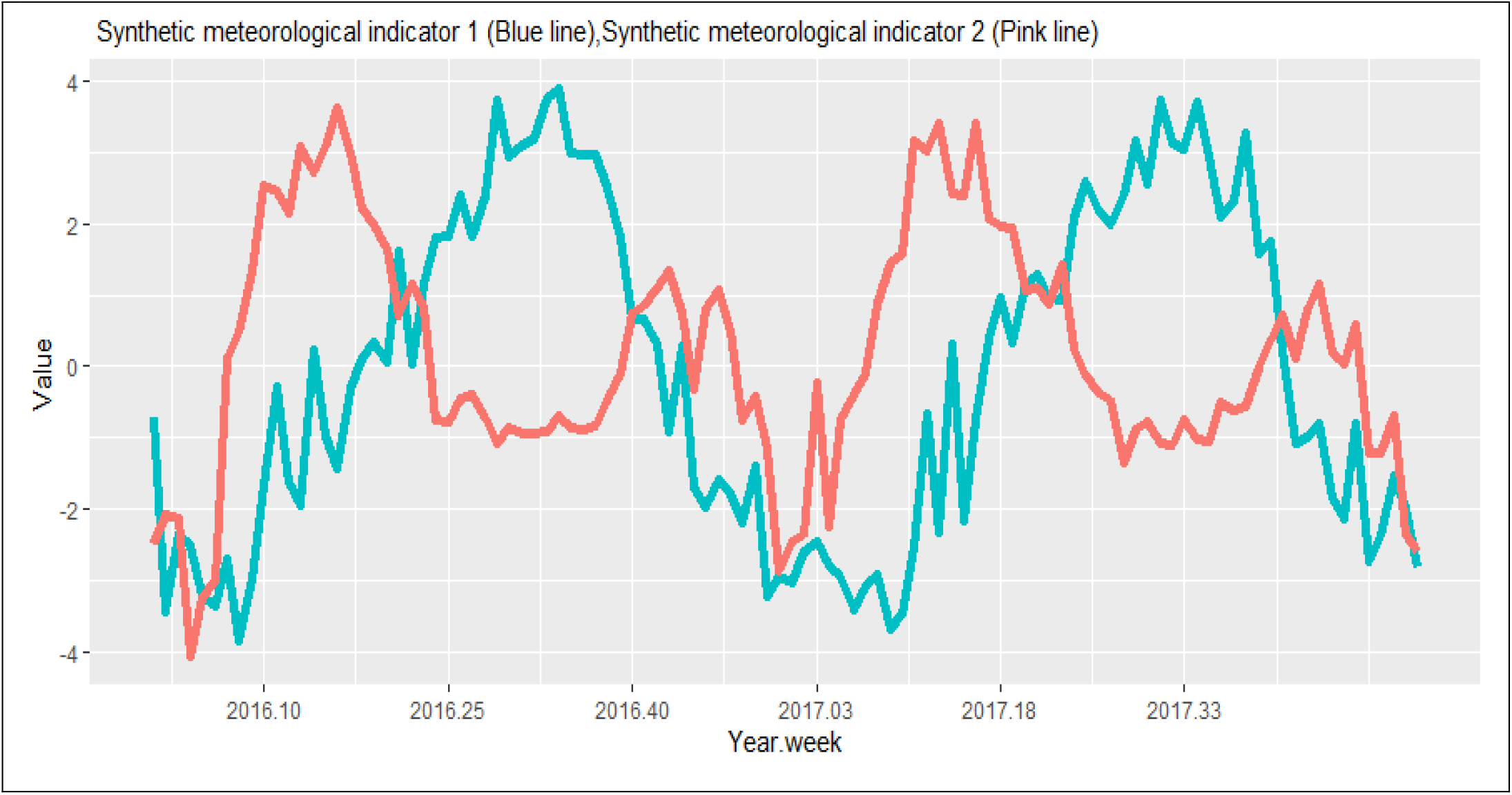
Time series of synthetic meteorological indicator 1 (mainly correlated with precipitation variables) and synthetic meteorological indicator 2 (mainly correlated with temperature variables) from 2016 to 2017.

### Spatial analysis

The spatial analysis allowed us to identify and map malaria hotspots for the epidemic year. A total of four hotspots were detected that accounted for 1,685 cases in 1,973 inhabitants, i.e. an average incidence rate of 854.02 cases per 1,000 person-years (Figure 5). These hotspots were mainly located in the southern and central parts of the study area. The hotspot with the highest risk (hotspot 1; relative risk (RR) = 4.06, p<0.0001) consisted of a single village (NIOMBOUNA) and accounted for 400 cases for 228 inhabitants, i.e. an incidence rate of 1,750.75 cases per 1,000 person-years. The second hotspot (hotspot 2; RR = 1.84, p<0.0001) was made up of three villages (SINKIRO, YELBELELA, and DANGBARA) and accounted for 604 cases for 753 inhabitants, i.e. an incidence rate of 802.12 cases per 1,000 person-years. The third hotspot (hotspot 3; RR = 1.92, p<0.0001) was made up of a single village (NIOMBRIPO) and accounted for 326 cases for 376 inhabitants, i.e. an incidence rate of 867.02 cases per 1,000 person-years. The fourth hotspot (hotspot 4; RR = 1.24, p=0.04) consisted of two villages (BOHERO and KPALBALO) and accounted for 355 cases for 616 inhabitants, i.e. an incidence rate of 576.2 cases per 1,000 person-years.

**Figure 5:**
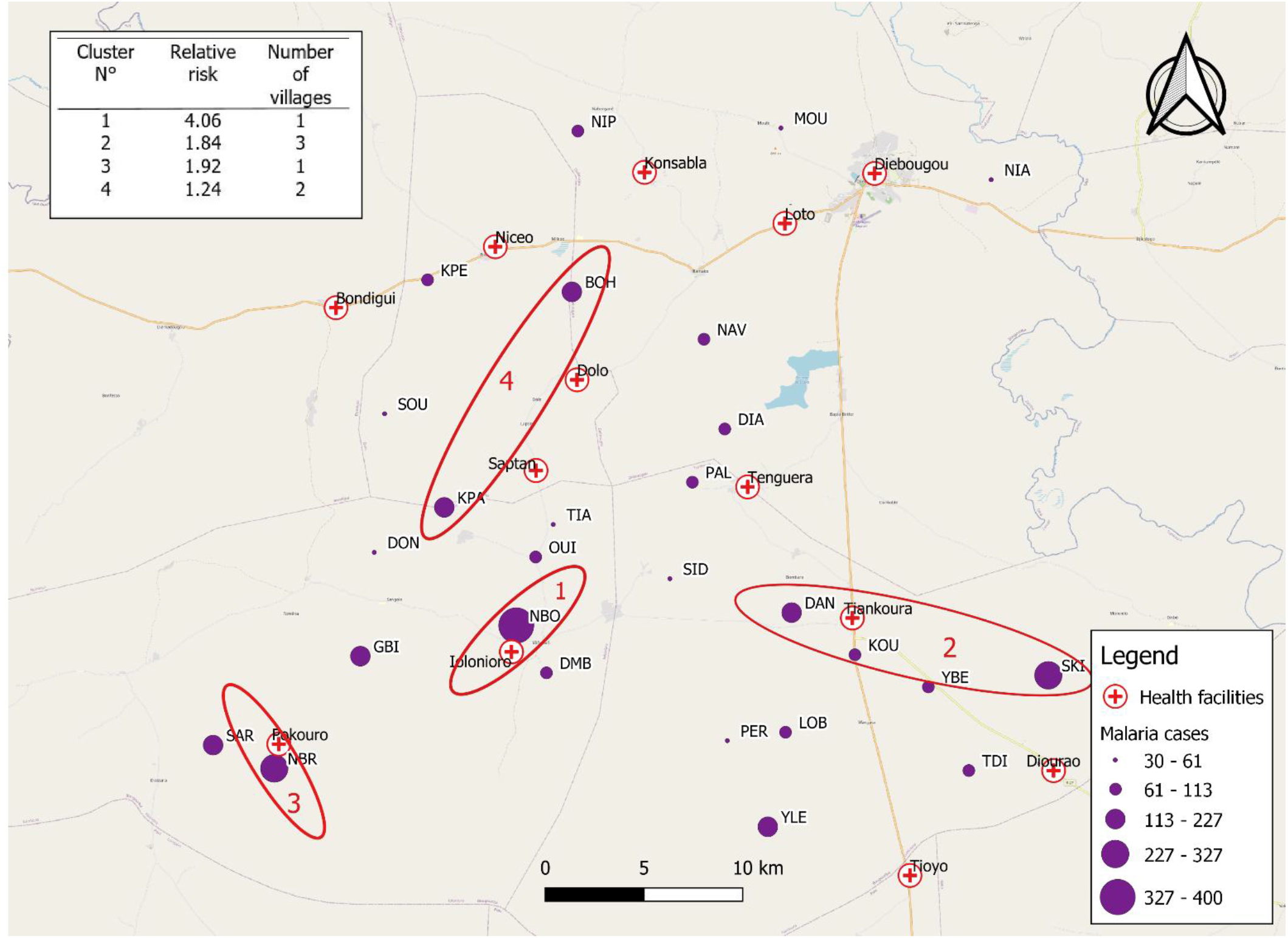
Map of malaria cases detected in 27 villages of Diébougou health district, Burkina Faso, for the epidemic year 2016-2017; hotspots identified with the Kulldorf scanning method. *Background: OpenStreetMaps*

### Temporal analysis

The time lag that generated the model with the lowest UBRE was 9 weeks for SMI1 and 16 weeks for SMI2.

The multivariate analysis found greater variability in incidence between HCs (standard deviation (SD) = 5.74) than between villages linked to the same HC (SD = 0.69). The coefficient of the temporal autocorrelation structure indicated the presence of temporal autocorrelation between cases (Phi = 0.32, 95% CI [0.20,0.38]).

In the multivariate model, lagged SMI1 and lagged SMI2 were significantly associated with the number of malaria cases at the village level (p<0.001 and p<0.001, respectively). The relationship between the number of cases and SMI1 (consisting mainly of precipitation variables) was positive and almost linear (Figure 6A) across the range of values. A positive non-linear relationship was observed for SMI2 (consisting mainly of temperature variables), with the number of cases increasing for SMI2 values above zero (Figure 6C). Below zero, changes in SMI2 values did not influence the number of cases (Figure 6C). The Euclidean distance between villages and their corresponding HCs was not correlated to the recorded malaria incidence (p=0.78).

**Figure 6:**
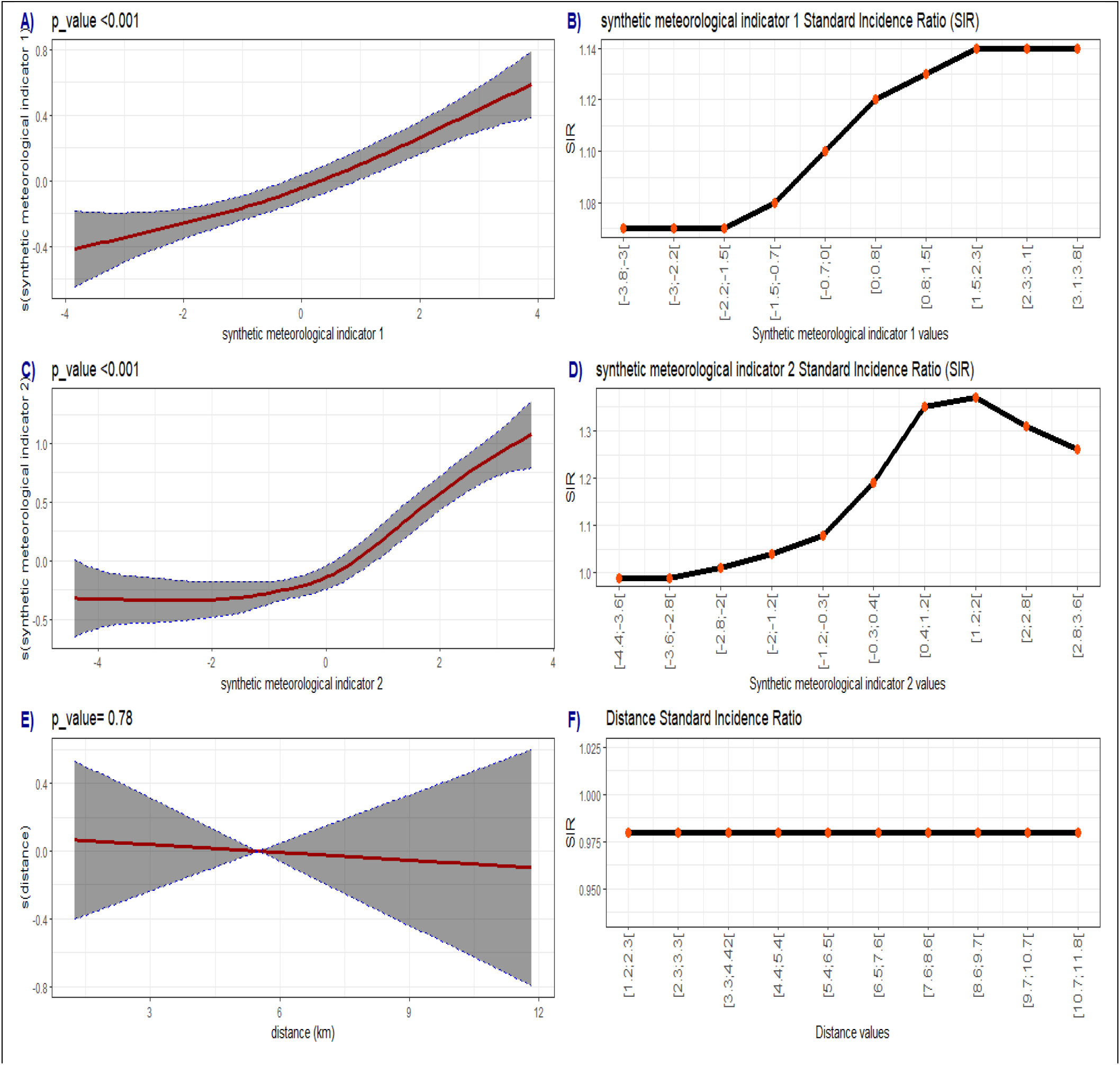
Relationship (red curve) between malaria cases and SMI1 (A), SMI2 (C), and Euclidean distance to health center (E) with 95% confidence intervals (shaded area). Evolution of the standard incidence ratio (SIR) as a function of SMI1 (mainly correlated with precipitation variables) (B), SMI2 (mainly correlated with temperature variables) (D), and Euclidean distance to health center (F).

The evolution of SIRs as a function of SMI values is presented in Figure 6. For SMI1, risk was constant over deciles 1 to 3 (SIR = 1.07, 95% CI [1.03, 1.10], [1.05, 1.08], and [1.06, 1.08], respectively), increased from decile 4 to 7, and then reached a plateau from decile 8 to 10 (SIR = 1.14 [1.14, 1.14]) (Figure 6B). For SMI2, risk was constant over deciles 1 to 2 (SIR = 0.99, 95% CI [0.93,1.05], and [0.98,1.00], respectively), increased from decile 4 to 8 (SIR = 1.37 [1.36,1.37]), and then decreased from decile 9 to 10 (Figure 6D).

### Prediction

The multivariate model generated for the epidemiological year was used to predict the number of cases in the 27 villages for all of 2016 and for the first 36 weeks of 2017. The resulting prediction was superimposed on the time series of reported cases for graphical analysis (Figure 7). The prediction followed the overall pattern of the time series of reported cases but with a tendency for underestimation, especially during the second peak in early 2017. In addition, the model predicted the onset of the malaria outbreak for the 2017-2018 epidemic year with a delay of three weeks.

**Figure 7:**
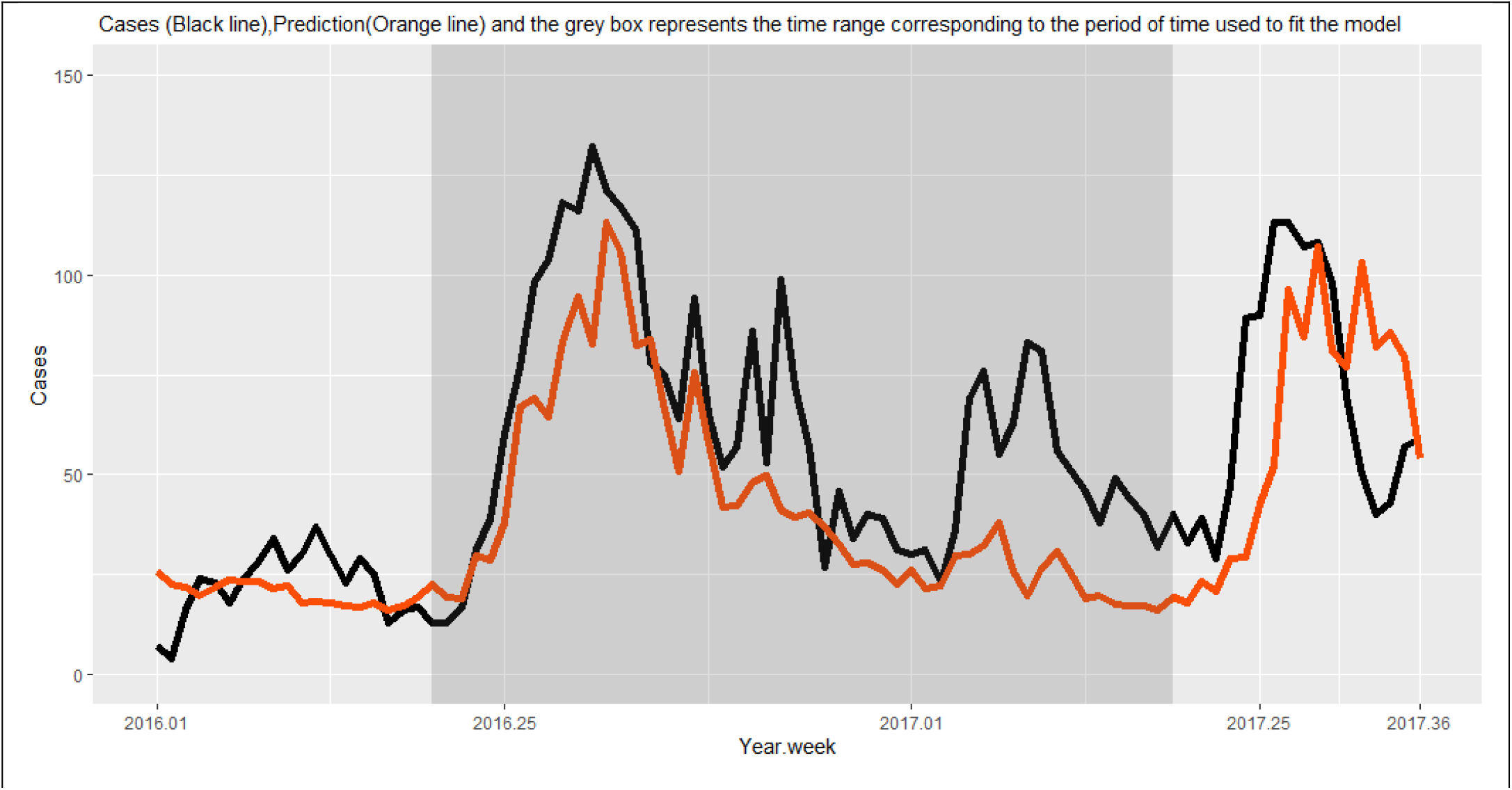
Cumulative number of reported cases (black line) and predicted cases (orange line) in 27 villages of South-Western Burkina Faso using a meteorological model.

## Discussion

In this study, we analyzed the spatio-temporal distribution of malaria cases in 27 villages of South-Western Burkina Faso.

The spatial analysis conducted using the Kulldorf scanning method helped to identify four malaria hotspots. The first three hotspots were located in the southern part of the study area and the last one was located in the central part, reflecting spatial heterogeneity in the distribution of cases. A comparison of the spatial distribution of these hotspots with that of mosquito vector density [37] showed no correlation between the two, leading us to conclude that the spatial heterogeneity of vector density does not explain the distribution of hotspots in our study area.

A number of studies have found an association between spatial inequalities in access to care and spatial heterogeneity of malaria incidence [42,43]. Yet, contrary to what has been reported elsewhere [44], we failed to found a correlation between the number of malaria cases and the Euclidean distance between villages and their corresponding HC. We used Euclidean distance because it is considered to be the simplest proxy for travel time, which is considered a good measure of access to care. However, Euclidean distance may not have been the best option, as roads in our study area are in highly variable condition and some become impassable during the rainy season, with some villages left completely isolated. Future studies in the region should use better proxys for travel time in trying to explain the detected hotspots [45].

Since entomological factors and spatial inequalities in access to care failed to explain the distribution of hotspots in our study area, other potential explanatory factors should be investigated in the future, including socio-economic factors (level of education, income, professional activity, individual and societal behavior, etc.) [38,39,40,41] and factors linked to LLIN usage [46,47,48,49]. Such investigations could help to explain in particular why the two hotspots composed of a single village (Niombripo and Niombouna) had a much higher incidence than neighboring villages. Nevertheless, hotspot analyses like ours make it possible to identify, in a simple and cost-efficient manner, villages that can constitute priority areas for intervention. Indeed, studies conducted elsewhere have shown that targeting hotspots helps to reduce malaria transmission [50,51]. This strategy is appropriate in resource-limited countries like Burkina Faso as it allows for efficient allocation of prevention resources [25,26].

Our analysis of the temporal dynamics of malaria cases found a strong correlation between malaria incidence and two SMIs with specific time lags. These SMIs were constructed through a PCA of meteorological data derived from readily and rapidly available satellite imagery. The first SMI (SMI1: positively correlated with cumulative rainfall, humidity, cloud cover, and number of rainy days, and negatively correlated with thermal amplitude) corresponded to the rainy season, while the second (SMI2: positively correlated with temperature and negatively correlated with atmospheric pressure) corresponded to the warm periods preceding and following the rainy season. We found that SMI1 and SMI2 predicted the number of cases with a time lag of 9 and 16 weeks, respectively, which is consistent with studies carried out in Burkina Faso, Mali, and Ethiopia [24,52,53].

In our study, the relationship between rainfall (SMI1) and the number of cases was quasi-linear, as was the case in a study performed in the Ouagadougou area of Burkina Faso [24]. By contrast, two studies conducted in the Sahel region – one in Mali (Niger River Valley, Timbuktu region) and the other in Senegal (Bambey and Fatick Health Districts) – found a monotonic non-linear relationship between rainfall and malaria incidence [25,26]. The drop in the number of cases above a certain level of cumulative rainfall observed in Mali and Senegal may be explained by the flushing out of larval breeding sites, which can lead to high mortality in Anopheles larval populations [54,55] and can reduce the human biting rate [55]. Vector populations are almost monospecific in these two countries: They are largely dominated by *An. Arabiensis* in Senegal [56,57] and by *An. coluzzii* in Mali [58]. These two species are also present in our study area and in the Ouagadougou area of Burkina Faso [59]. However, in both these areas, they live in sympatry with both *An. gambiae s*.*s*. and *An. funestus* [37,60,61,62]. The quasi-linear relationship observed in our study between rainfall and the number of malaria cases may be explained by the fact that these species are not very susceptible to flushing out, due to rapid larval development in the case of *An. gambiae s*.*s*. [63,64] and to a preference for deeper environments in the case of *An. funestus* [65]. These species may therefore relay *An. coluzzii* when abundances of this later fall due to excessive rainfall.

In our study, the relationship between the number of malaria cases and temperature (SMI2) was non-linear. This is consistent with findings from two other studies conducted in the Sahel region (in Mali and in the Ouagadougou area of Burkina Faso) [24,25]. However, unlike these studies, we found no negative relationship between the number of malaria cases and temperature at higher temperature values. This discrepancy may be explained by the fact that temperatures can reach higher values in Mali and in the Ouagadougou area (>34°C) than in the Diébougou region, which is sufficient to inhibit the development of Anopheles larvae [66] and to reduce the survival of adult Anopheles [67,68]. In addition, we found that below a certain temperature, an increase in temperature had no effect on the number of cases (a finding also observed by Cissoko et al. [16]). Our hypothesis is that the increase in temperature, which should favor the development of Anopheles, is compensated by another phenomenon at low SMI2 values. While this phenomenon has yet to be clearly identified, high levels of LLIN usage during cooler periods may be a contributing factor [47].

Our spatio-temporal model fitted with two lagged SMIs and case data for a single epidemiological year helped to predict the start of the next outbreak nine weeks in advance, but with an error of three weeks (i.e. the actual outbreak began three weeks before the prediction). The prediction was good enough to make it possible to issue early warnings and to organize local prevention campaigns ahead of time. Our model could probably be improved with routine inclusion of new data and regular updated predictions. For this purpose, data from HC consultations should be made available quickly, ideally at the same pace as ERA5 meteorological data (i.e. within five days). This can easily be achieved by using connected tablets for data entry.

## Conclusion

In this study, a spatial analysis was conducted that highlighted the spatial heterogeneity of malaria cases and helped to identify four malaria hotspots in South-Western Burkina Faso. In the temporal analysis, an effective predictive model was built with data obtained through passive case detection and with simple and accessible meteorological data. Future studies should further investigate the detected hotspots to identify the local determinants of transmission. Our spatio-temporal analysis provides a powerful prospective method to identify high-risk areas that may constitute priority areas during malaria prevention campaigns.

## Data Availability

The datasets analyzed in this study may be available from the last author on reasonable request.

## Acronyms and Abbreviations

WHO: World Health Organization
CI: Confidence Interval
IPT: Intermittent Preventive Treatment
RDT: Rapid Diagnostic Test
SMC: Seasonal Malaria Chemoprevention
LLIN: Long-Lasting Insecticidal Net
IRS: Indoor Residual Spraying
HC: Health Center
SMI: Synthetic Meteorological Indicator
PCA: Principal Component Analysis
GAM: Generalized Additive Model
GLM: Generalized Linear Model
UBRE: Unbiased Risk Estimator
GAMM: Generalized Additive Mixed Model
SIR: Standardized Incidence Ratio
RR: Relative Risk
SD: Standard Deviation

## Ethics approval and consent to participate

The protocol of this study was reviewed and approved by the Institutional Ethics Committee of the Institut de Recherche en Sciences de la Santé (IEC-IRSS) and registered as N°A06/2016/CEIRES.

## Consent for publication

Not applicable

## Availability of data and materials

The datasets analyzed in this study may be available from the last author on reasonable request.

## Competing interests

The authors declare that they have no competing interests

## Funding

This work was part of the REACT project, funded by the French Initiative 5% – Expertise France (No. 15SANIN213). C.S.B. received a grant from the *Revivre Développement* Endowment Fund through the NGO *Prospective et Coopération*, as well as a French Government Grant through the French Embassy in Burkina Faso.

The funders had no role in study design, data collection and analysis, decision to publish, or preparation of the manuscript.

## Contributions

C.S.B., J.G., CP, AK, RKD and N.M. designed the study; A.S. and D.D.S. conducted the field study; C.S.B., J.G., and N.M. designed the statistical analysis plan; C.S.B. conducted the statistical analysis under the supervision of J.G. and N.M.; C.S.B. performed the cartographic analysis with the participation of M.C. and P.T.; S.D. participated in the statistical analysis; C.S.B, J.G., and N.M. validated and interpreted the results; C.S.B., J.G., and N.M. wrote the manuscript; all authors read and approved the final manuscript.

## Acknowledgements

The authors would like to thank the Ministry of Health of Burkina Faso, in particular Dr. Dembélé Henri, Médecin–chef de poste in Diebougou, and the local medical team, for facilitating data collection. Special thanks are due to Mr. Maiga Issouf for his strong involvement in data collection. We are also grateful to the “*Laboratoire Mixte International sur les Maladies à Vecteurs*” (LAMIVECT) for providing technical support. Lastly, we would like to thank Mr. Ouattara Adama and Mr. Félix Zoumènou for providing administrative support.

## Notes

### Competing Interest Statement

The authors have declared no competing interest.

### Author Declarations

The protocol of this study was reviewed and approved by the Institutional Ethics Committee of the Institut de Recherche en Sciences de la Santé (IEC-IRSS) and registered as # A06/2016/CEIRES.

